# Risk Factors Analysis of Acute Kidney Injury in Adult Patients Receiving Extracorporeal Membrane Oxygenation

**DOI:** 10.1101/2020.04.06.20055145

**Authors:** Zhixiang Mou

**Author notes:** **Funding:** This research received no external funding. **Conflicts of Interest:** The authors declare no conflict of interest.

## Abstract

**Background:** Acute kidney injury (AKI) has been reported as one of the most common complication in patients receiving extracorporeal membrane oxygenation (ECMO), the risk factors of AKI on ECMO is unknown. This meta-analysis aimed to find out the risk factors of AKI among adult patients receiving ECMO.

**Methods:** A literature search was performed using PubMed,Web of Science, and Embase fulfilled the pre-specified criteria until April 2020 to include studies reported the necessary clinic characteristics, then the gender, cancer, diffuse intravascular coagulation (DIC), massive/severe Bleeding, intra-aortic balloon pump (IABP), post-cardiotomy, diabetes mellitus, liver cirrhosis and ECMO support duration were pooled for further analysis by STATA to get conclusion.

**Results:** This research is first time to provide the evidence that patients developed AKI/severe AKI with a longer ECMO support duration (pooled WMD, 4.09 days; 95% CI: 2.45-5.73 days, *Z*= 4.89, *P*= 0.000, *I*^2^= 73.4%, χ^2^*P*= 0.023/pooled WMD, 1.52 days; 95% CI: 0.19-2.85 days, *Z*= 2.25, *P*= 0.025, *I*^2^= 77.4%, χ^2^*P*= 0.001) and the risk of severe AKI requiring RRT was higher (pooled OR, 2.22; 95% CI: 1.24-3.99, *Z*= 2.68, *P*= 0.007, *I*^2^= 0.0%, χ^2^*P*= 0.634) in liver cirrhosis patients by systemic analysis, indicated ECMO support duration and liver cirrhosis may act as risk factors of AKI in adult patients received ECMO.

## 1. Introduction

Extracorporeal membrane oxygenation (ECMO) is a temporary support system for critical patients with life-threatening cardiac and/or respiratory cardiac failure, especially have failed conventional management (1). It is a process drains blood from the venous system, pumps the blood through an oxygenator providing gas exchange, and returns the oxygenated blood to the venous (VV) or arterial (VA) circulation (2). The indications for ECMO therapy are beyond the scope compared to 40 years ago and still keep evolving due to the progress in the ECMO equipment design and technology improvement, it is administered for treating patients with severe reversible cardiac dysfunction (acute cardiogenic shock, cardiac arrest, cardiac failure or postcardiac surgery), any potentially reversible acute respiratory failure associated with/without pneumonia (viral or bacterial), a short-term bridge to transplantation or mechanical support, awaiting recovery of organ function or a definitive therapy may effective (3–7). Despite the improvement in survival using ECMO, acute kidney injury (AKI) is the most frequently reported problem among a number of complications (8,9) may cover the potential benefit of ECMO, and associated with poorer prognosis and increased mortality (10–12). There are insufficient mechanism and clinical data defining the link between ECMO and AKI, a comprehensive understanding of this relationship would help us to predict, treat, and improve the clinical outcomes. Previously studies focused on the incidence and mortality of AKI among patients receiving ECMO, or discussed the factors influencing AKI patients survival on ECMO. However, it is lack of systematic analysis for the risk factors of AKI in adult patients receiving ECMO; in addition, the risk factors of severe AKI among adult patients receiving ECMO also remains unclear.

The aim of this meta-analysis is to find out the risk factors of AKI and severe AKI in adult patients receiving ECMO by pooling the available data from publications.

## 2. Materials and Methods

### 2.1. Search Strategy

The protocol for this meta-analysis was registered with International Prospective Register of Systematic Reviews (being assessed, PROSPERO ID. 177109). A systematic literature search was performed through PubMed, Web of Science, and Embase utilizing the search terms “acute kidney” OR “acute renal” AND “extracorporeal membrane oxygenation” OR “ECMO” OR “extracorporeal life support” OR “ECLS” until March 2020 to summarize the risk factors of AKI among adult patients on ECMO. Each study was evaluated for inclusion or exclusion for this analysis. No language or date restriction executed. A manual search for references of included articles was also performed. The meta-analysis was conducted and handled according to the guidelines of the Preferred Reporting Items for Systematic Reviews and Meta-Analyses (PRISMA). (http://www.prismastatement.org/).

### 2.2. Inclusion/Exclusion Criteria

For the purpose of this analysis, eligible included studies reported the risk factors of AKI in adult patients underwent ECMO and met the Population, Interventions, Comparison and Outcomes (PICO) criteria.

All the selected studies fulfilled the following criteria: original case-control studies provided the data of AKI requiring/not requiring renal replacement therapy (RRT) among adult patients (age ≥ 18 years) during ECMO with consideration of intervention factors; reported the specific indications on ECMO regardless of ECMO types; reported comprehensive clinic characteristics of AKI patients on ECMO regardless of AKI defined; reported comprehensive clinic characteristics of RRT patients receiving ECMO regardless implantation modality; with history of kidney diseases pre-ECMO patients while can not distinguished after receiving ECMO were excluded; case/case series reports containing< 10 patients were excluded; duplicate articles were excluded; no restriction on language/year applied in full text written.

### 2.3. Data Extraction and Study Quality

A data collecting spreadsheet (Microsoft Corporation, Redmond, WA) was established to extract the following data from each included study: the first author, publication year, region, study design, number of patients, AKI/severe AKI definition, risk factors of AKI/severe AKI related ECMO. No attempt to obtain specific or missing data from the authors.

All included studies were assessed for the quality using the Newcastle-Ottawa Quality Assessment Scale (NOS) containing 3 aspects (selection, comparability and exposure) and 8 items (13). It ranges from 0 to 9 scores, studies with a score≧6 were considered to meet adequate methodological quality.

### 2.4. AKI and severe AKI definitions

RIFLE (the Risk of renal failure, Injury to the kidney, Failure of kidney function, Loss of kidney function and End stage kidney disease), AKIN (the Acute Kidney Injury Network), and KDIGO (the Kidney Disease Improving Global Outcomes) were the three major criteria to define AKI (14–15). In this meta-analysis, AKI definition is clear once the record has defined according to one of the guidelines; severe AKI definition is that record discriminated AKI by RRT regardless modality, AKI severity stage RIFLE-F, KDIGO stage 3 and AKIN stage need RRT.

### 2.5. risk factors related AKI

All possible risk factors potentially effect renal function during ECMO should be identifed and screened. However, only gender, cancer, diffuse intravascular coagulation (DIC), massive/severe bleeding, intra-aortic balloon pump (IABP), post-cardiotomy, diabetes mellitus, liver cirrhosis and ECMO support duration were pooled for further analysis due to insufficient data and inconsistent AKI definition.

### 2.6. Statistical Analysis

Statistical analysis was performed by STATA statistical software (version 14.0; Stata Corp, College Station, TX). In the analysis, dichotomous data were analyzed using random effects models and Mantel-Haenszel method, continuous data were analyzed using random effects models and inverse-variance method; *I*^2^ test was used to evaluate heterogeneity due to probability variance between studies; the pooled odds ratio (OR), weighted mean difference (WMD) and their corresponding 95% confidence intervals (CI) were used to evaluate the relationship between risk factors and AKI/severe AKI; *Z* test was used to assess the significance of the pooled ORs and WMDs; graphically plotted using forest plots to perform the treatment effects and heterogeneity. χ^2^*P* value< 0.05 or *I*^2^ test> 50% was defined as statistically significant heterogeneity; *Z* test *P* value< 0.05 was considered statistically significant.

## 3. Results

### 3.1 Literature Search and Study Quality Assessment

The flow diagram of the studies with selection procedure and reasons for exclusion was shown in detail in Figure 1. A total of 4416 potentially eligible articles were identified from 3 databases (PubMed, n= 1445; Web of Science, n= 617; Embase, n= 2354) with the search approach until April, 2020. After excluding duplicates (2000 records), 2416 titles and abstracts were screened manually for eligibility. Then in-vitro studies, focused on pediatric or neonate population, animal studies, case reports, review articles and congress abstracts were excluded, 80 remaining articles were full-length reviewed for eligibility. The studies contained< 10 patients or unmatched data were also excluded. 9 original case-control studies involving 4193 patients fulfilled the pre-specified criteria were included for the meta-analysis (Table 1). Six studies (66.7%) with a score≧6 were considered good quality according to the NOS criteria (Table 2).

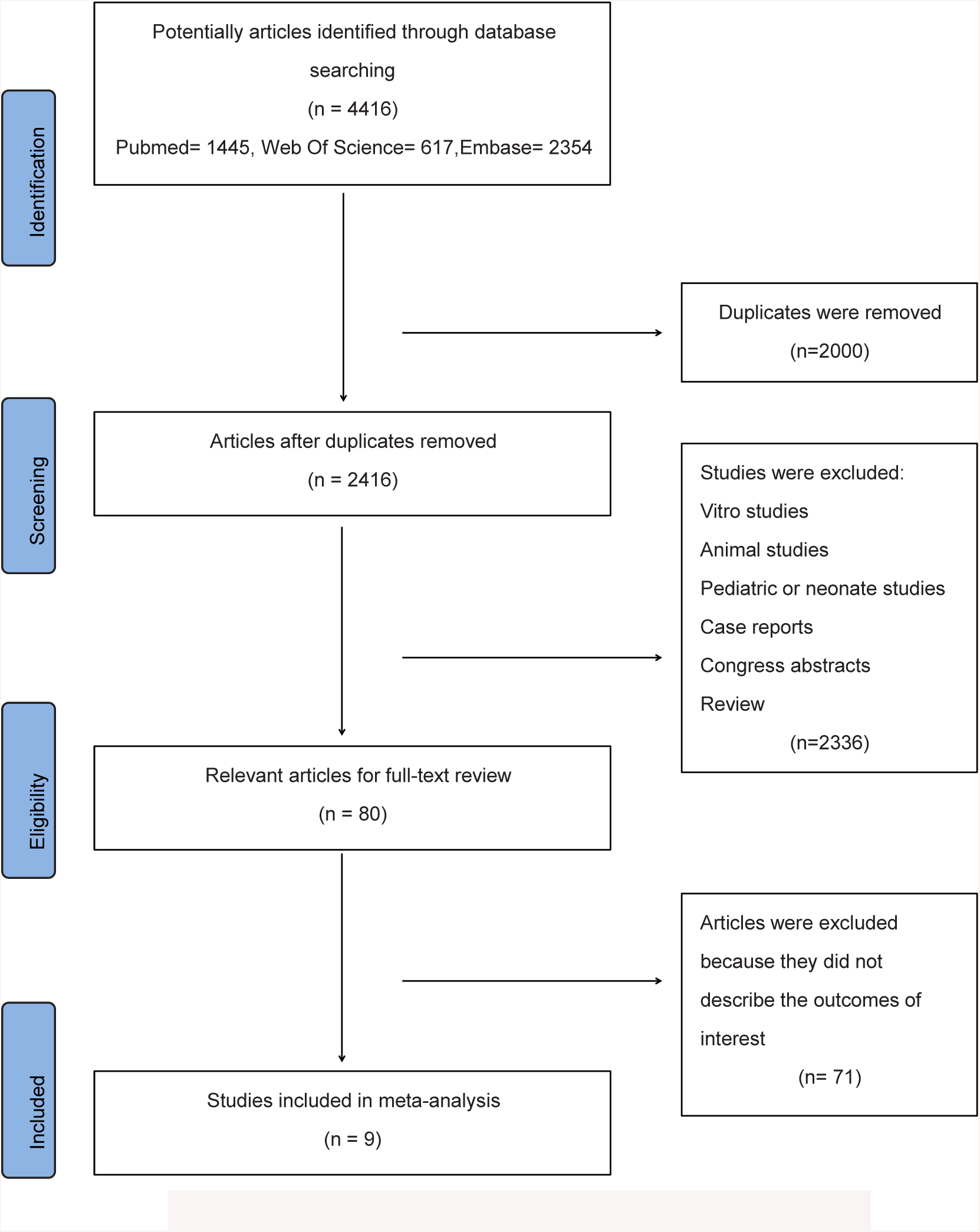

**Table 1.** study characteristics

| Authors | year | region | study design | risk factors for analysis | No.of patients | AKI Definition | ECMO modes |
| --- | --- | --- | --- | --- | --- | --- | --- |
| Lin (16) | 2006 | Taiwan | case-control | Gender, ECMO support duration | 46 | RIFLE | N/A |
| Lee (17) | 2015 | Korea | case-control | gender, cancer, IABP, ECMO support duration | 322 | KDIGO | VA ECMO/VV ECMO |
| Antonucci (18) | 2016 | Belgium | case-control | gender, cancer, DIC, massive bleeding, DM, liver cirrhosis,ECMO support duration | 135 | AKIN | VA ECMO/VV ECMO |
| Lyu (19) | 2016 | China | case-control | gender, IABP,ECMO support duration | 84 | RIFLE | VA ECMO |
| Delmas (20) | 2017 | France | case-control | Gender, ECMO support duration | 60 | KDIGO | VV ECMO |
| Chang (21) | 2017 | China | case-control | gender, DIC, massive bleeding, post-cardiotomy, ECMO support duration | 71 | AKIN | VA ECMO/VV ECMO |
| Devasagayaraj (22) | 2018 | USA | case-control | gender, massive bleeding, ECMO support duration | 54 | RRT | VV ECMO |
| Liao (23) | 2018 | China | case-control | gender, IABP, post-cardiotomy, DM, ECMO support duration | 170 | KDIGO | VA ECMO/VV ECMO |
| Chen (24) | 2019 | Taiwan | case-control | gender, cancer, IABP, post-cardiotomy, DM, liver cirrhosis, ECMO support duration | 3251 | RRT | N/A |

**Table 2.**
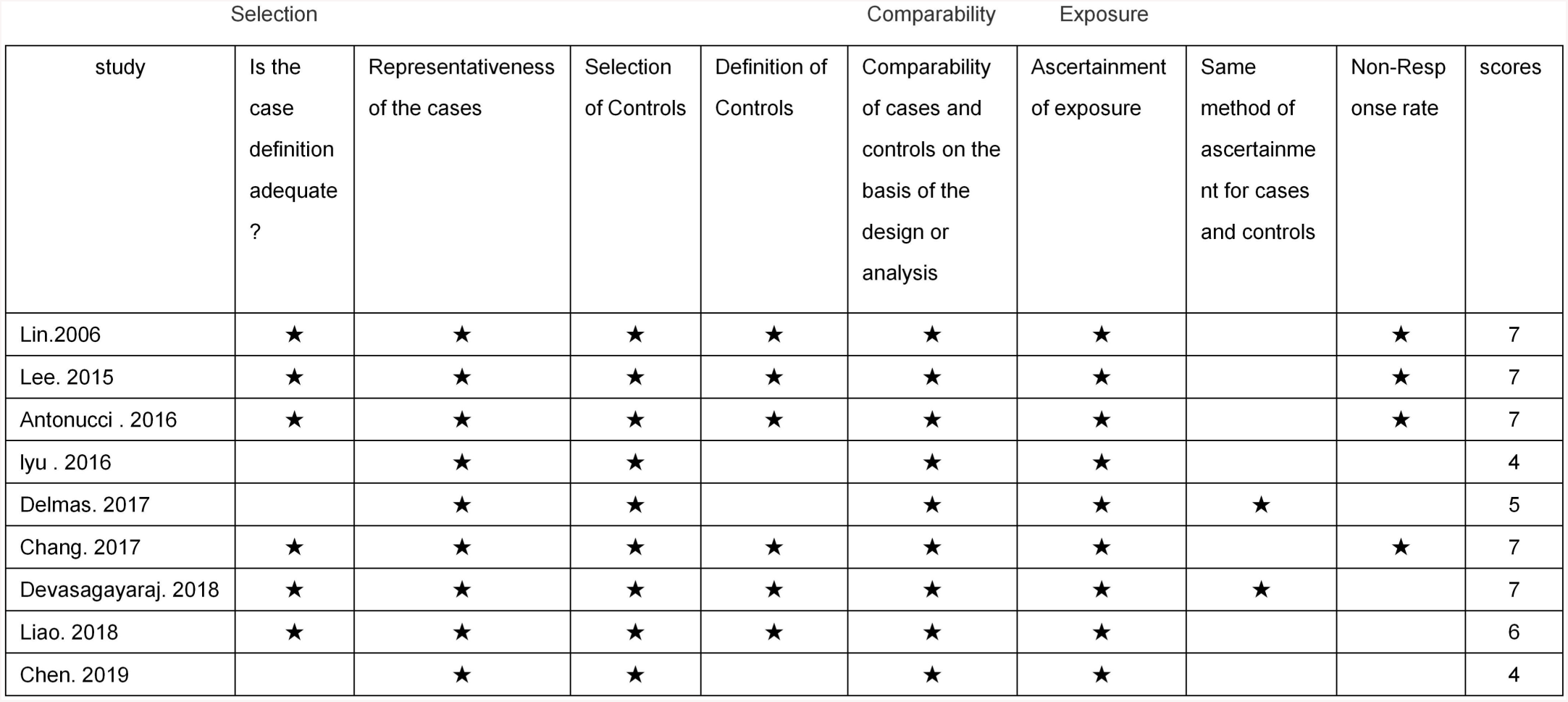
Results of quality assessment using Newcastle-Ottawa Scale for case-control studies

**Figure 1.**
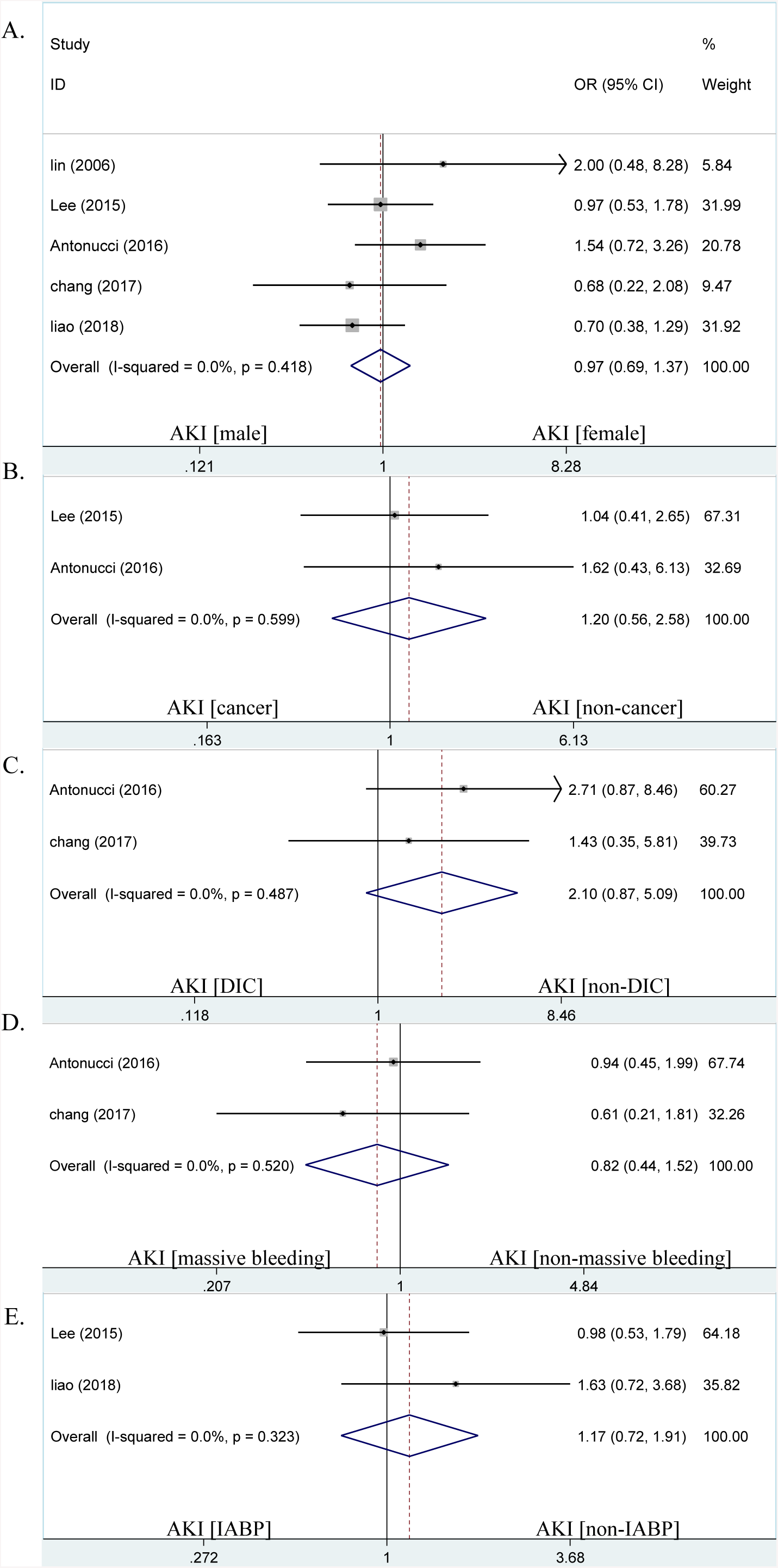
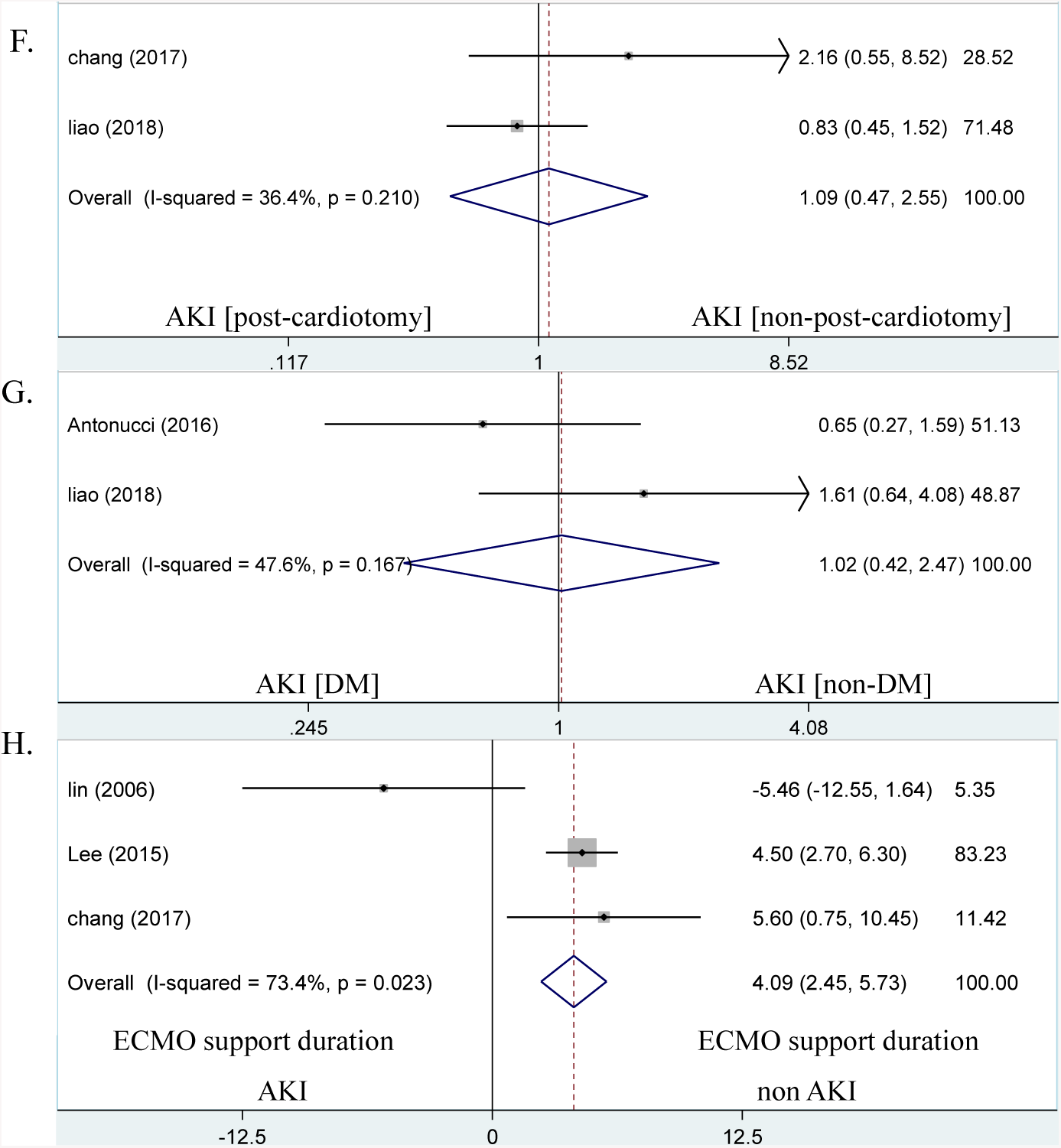
Forest plots of the included studies assessing risk factors of AKI in ECMO patients. The *solid vertical line* indicates no effect. The *horizontal lines* represent the 95% confidence interval (CI). (A) supposing gender as risk factor and analysis; (B) supposing cancer as risk factor and analysis; (C) supposing DIC as risk factor and analysis; (D) supposing massive/severe bleeding as risk factor and analysis; (E) supposing IABP as risk factor and analysis; (F) supposing post-cardiotomy as risk factor and analysis; (G) supposing DM as risk factor and analysis; (H) supposing ECMO support duration as risk factor and analysis.

### 3.2 Study Characteristics

In this meta-analysis, seven studies (77.8%) provided the data with defined AKI/severe AKI according to the one of guidelines proposed by RIFLE, AKIN, or KDIGO. Two studies (22.2%) provided the data with defined severe AKI due to RRT. 4193 patients receiving ECMO treatment between April 2002 and May 2016 were identified and record supporting time as being eligible for analysis. Of these patients, 69.8% (n= 2926) male and 30.2% (n= 1267) female received ECMO; 2144 patients received ECMO due to post-cardiotomy in three studies; three studies reported 184 patients on ECMO with cancer; two studies reported 40 patients experienced DIC during ECMO; three studies record 112 patients suffered severe bleeding; 2046 patients used IABP concomitantly with ECMO in four studies; three studies reported diabetes mellitus (DM) acted as comorbid in 893 ECMO patients, while two studies reported 57 ECMO patients with history of liver cirrhosis respectively (Table 1).

### 3.3 analysis risk factors of AKI in ECMO patients

A series meta-analysis were performed met the pre-defined criteria, unfortunately, the process was often accomplished only by two or three studies due to limited and inconsistent data described. There was no significant difference supposed gender as a risk factor of AKI (pooled OR, 0.97; 95% CI: 0.69-1.37, *Z*= 0.16, *P*= 0.870, Figure 1A) and heterogeneity difference was not observed (*I*^2^= 0.0%, χ^2^*P*= 0.418) among the eligible studies, indicating that gender is not a risk factor of AKI in ECMO patients.

The following analysis indicated that with cancer (pooled OR, 1.2; 95% CI: 0.56-2.58, *Z*= 0.48, *P*= 0.633, *I*^2^= 0.0%, χ^2^*P*= 0.599, Figure 1B), experienced DIC (pooled OR, 2.1; 95% CI: 0.87-5.09, *Z*= 1.65, *P*= 0.099, *I*^2^= 0.0%, χ^2^*P*= 0.487, Figure 1C), suffered massive/severe bleeding (pooled OR, 0.82; 95% CI: 0.44-1.52, *Z*= 0.63, *P*= 0.526, *I*^2^= 0.0%, χ^2^*P*= 0.52, Figure 1D), used IABP concomitantly (pooled OR, 1.17; 95% CI: 0.72-1.91, *Z*= 0.65, *P*= 0.52, *I*^2^= 0.0%, χ^2^*P*= 0.323, Figure 1E), post-cardiotomy (pooled OR, 1.09; 95% CI: 0.47-2.55, *Z*= 0.2, *P*= 0.841, *I*^2^= 36.4%, χ^2^*P*= 0.21, Figure 1F), and history of DM (pooled OR, 1.02; 95% CI: 0.42-2.47, *Z*= 0.04, *P*= 0.971, *I*^2^= 47.6%, χ^2^*P*= 0.167, Figure 1G) were also not risk factors of AKI in ECMO patients, all heterogeneity difference were not observed.

While, when the meta-analysis was exercised supposed ECMO supporting duration as a risk factor of AKI, the resulted showed that ECMO patients developed AKI with a longer ECMO support duration (pooled WMD, 4.09 days; 95% CI: 2.45-5.73 days, *Z*= 4.89, *P*= 0.000, *I*^2^= 73.4%, χ^2^*P*= 0.023, Figure 1H), indicated that ECMO support duration may act as a risk factor of AKI in ECMO patients. The original source of heterogeneity may come from the different definition of AKI.

### 3.3 analysis risk factors of severe AKI in ECMO patients

The following meta-analysis were performed to find out the risk factors of severe AKI among ECMO patients. There were no significant difference supposed gender (pooled OR, 0.96; 95% CI: 0.77-1.19, *I*^2^= 13.6%, χ^2^*P*= 0.326, *Z*= 0.40, *P*= 0.69, Figure 2A), with cancer (pooled OR, 1.01; 95% CI: 0.75-1.37, *Z*= 0.08, *P*= 0.938, *I*^2^= 0.0%, χ^2^*P*= 0.701, Figure 2B), used IABP concomitantly (pooled OR, 0.98; 95% CI: 0.59-1.64, *Z*= 0.07, *P*= 0.947, *I*^2^= 73.5%, χ^2^*P*= 0.023, Figure 2C), and history of DM (pooled OR, 1.12; 95% CI: 0.96-1.31, *Z*= 1.4, *P*= 0.162, *I*^2^= 0%, χ^2^*P*= 0.931, Figure 2D) as risk factors of severe AKI, heterogeneity difference were also not observed among the eligible studies (except IABP analysis), indicating these characteristics were still not risk factors of severe AKI in ECMO patients.

**Figure 2.**
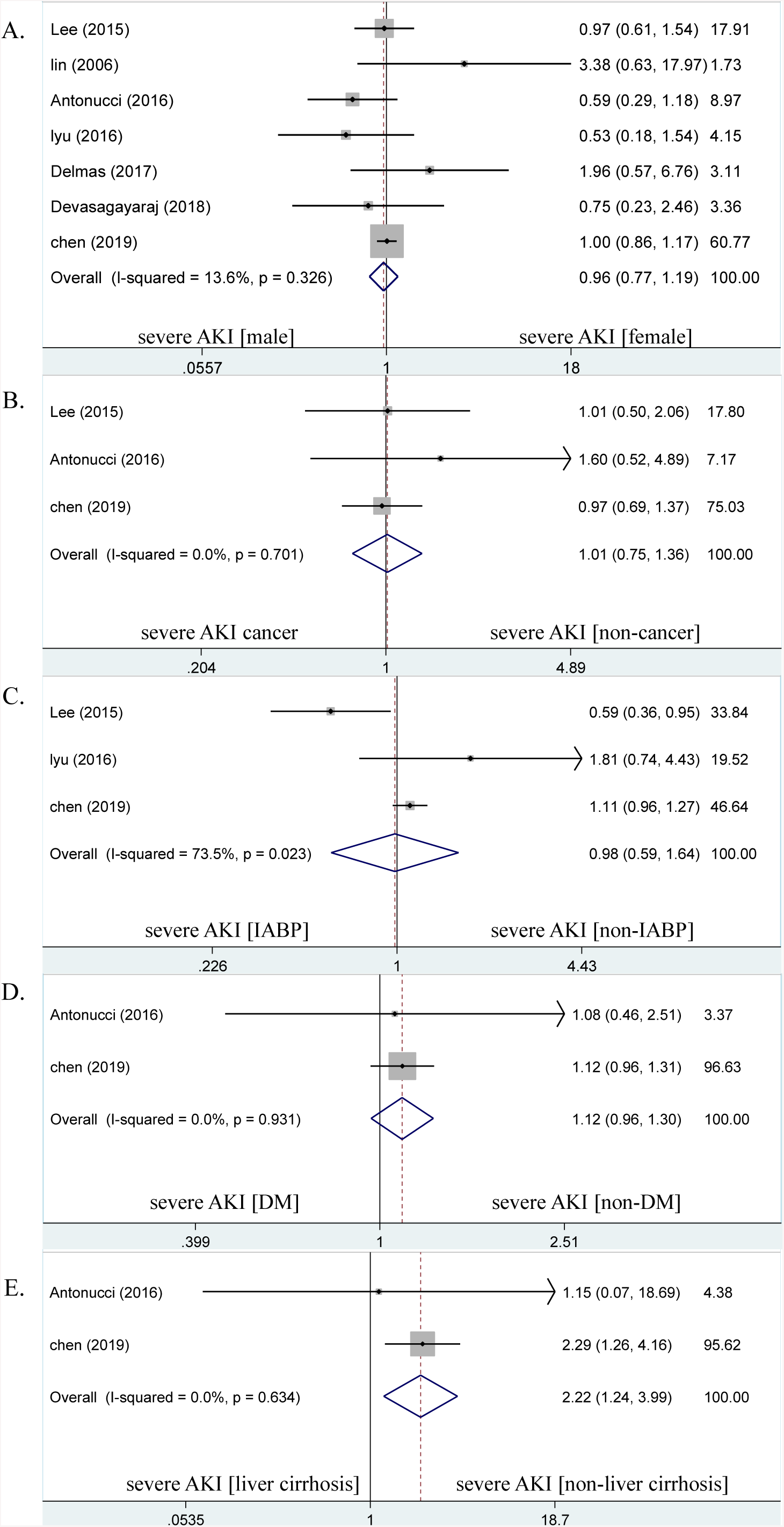
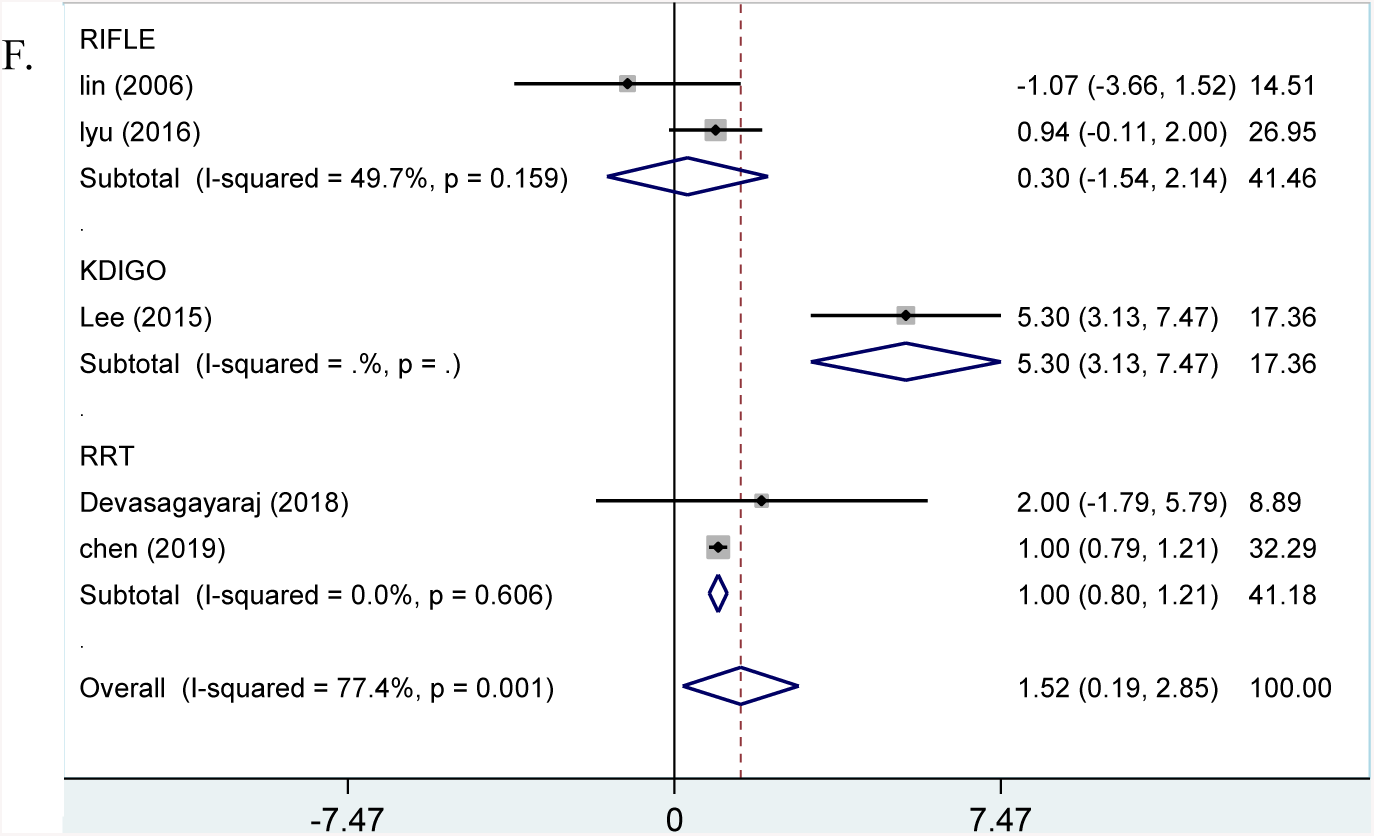
Forest plots of the included studies assessing risk factors of severe AKI in ECMO patients. The *solid vertical line* indicates no effect. The *horizontal lines* represent the 95% confidence interval (CI). (A) supposing gender as risk factor and analysis; (B) supposing cancer as risk factor and analysis; (C) supposing IABP as risk factor and analysis; (D) supposing DM as risk factor and analysis; (E) supposing liver cirrhosis as risk factor and analysis; (F) supposing ECMO support duration as risk factor and analysis.

While, when the meta-analysis was exercised supposed live cirrhosis as a risk factor of severe AKI, the resulted showed that the risk of severe AKI was estimated 2.22 times higher (pooled OR, 2.22; 95% CI: 1.24-3.99, *Z*= 2.68, *P*= 0.007, *I*^2^= 0.0%, χ^2^*P*= 0.634, Figure 2E) in liver cirrhosis patients group than non-liver cirrhosis patients group, indicated that liver cirrhosis may act as a risk factor of severe AKI in ECMO patients.

Consistent with the previous analysis, when the meta-analysis was performed supposed ECMO supporting duration as a risk factor of severe AKI, the result showed that ECMO patients also developed severe AKI with a longer ECMO support duration (pooled WMD, 1.52 days; 95% CI: 0.19-2.85 days, *Z*= 2.25, *P*= 0.025, *I*^2^= 77.4%, χ^2^*P*= 0.001, Figure 2F), indicated that ECMO support duration may act as a risk factor of severe AKI in ECMO patients either. Then subgroup analysis was performed to explore the original source of heterogeneity according to the definition of AKI. The result of RRT subgroup analysis (pooled WMD, 1.003 days; 95% CI: 0.8-1.21 days, *Z*= 9.53, *P*= 0, *I*^2^= 0%, χ^2^*P*= 0.606, Figure 2F) was consistent with the overall result; there were no significant difference in the RIFLE subgroup analysis (pooled WMD, 0.3 days; 95% CI: -1.54-2.14 days, *Z*= 0.32, *P*= 0.75, *I*^2^= 49.7%, χ^2^*P*= 0.159, Figure 2F). Subgroup analysis showed that significant heterogeneity observed perhaps due to inconsistent AKI definition.

## 4. Discussion

This population-based meta-analysis provided evidence that adult patients received ECMO developed AKI/severe AKI with a longer ECMO support duration, the risk of severe AKI in adult patients received ECMO with liver cirrhosis was higher than the patients received ECMO without liver cirrhosis, demonstrated that ECMO support duration and liver cirrhosis may be risk factors of AKI in adult patients received ECMO.

Rapidly increased implementation of ECMO in numerous critical care centers due to the improved the risk: beneft ratio (25,26). AKI develops during ECMO is frequently observed and associated with poorer outcomes and higher mortality (27–29). The pathophysiological mechanisms of AKI during ECMO remains poorly understood, it is complex, multifactorial, synergistic and involves in both patient and ECMO related factors (Figure 3). Activation of pro-inflammatory cytokines such as interleukin-1 (IL-1), IL-6, IL-8, and tumour necrosis factor-alpha (TNF-α) released from activated leukocytes due to primary disease process, combined with the interaction of blood and exposure of non-self ECMO membrane, may initiate and amplify a systemic inflammatory response leading to renal inflammation injury and microcirculation dysfunction (26,28,30–34). Additionally, ECMO limited pulsatility, variations in ECMO circuit pressures, renal hypoperfusion from damaged cardiac function, diuretic therapy and inotrope/vasopressor administration may contribute to the hemodynamic instability and renal microcirculation dysfunction, finally lead to reduced glomerular filtration pressure and decreased GFR (26,28,34–36).

**Figure 3.**
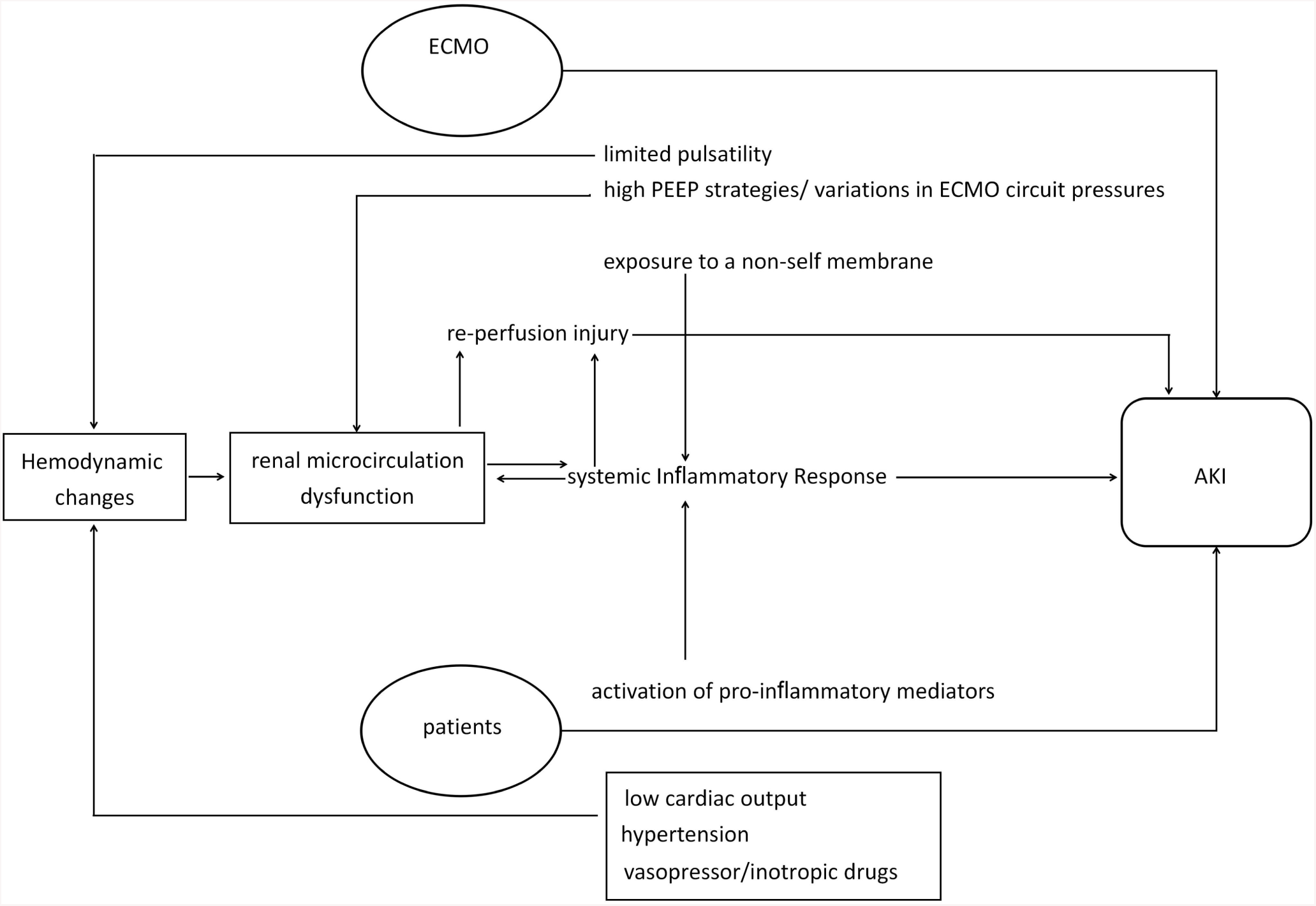
potentially mechanisms of AKI on ECMO patients

One of this meta-analysis result is similar with a retrospective cohort study conducted by Dr. Kielstein, they demonstrated that the percentage of RRT required after ECMO implantation growing as the duration of ECMO support last (12). Since the duration of ECMO increased, the pro-inflammatory cytokine release and adhesion of hemocytes to the endothelial vessel lining are also increased, progressively increase the risk of AKI development; prolonged poor perfusion with limited pulsatile blood through renal proximal tubules would break off sodium transport, lead to cell wall destruction, cell swelling and cell death (37). Furthermore, haemolysis and sub-haemolytic damage induced by the shear stress of the ECMO circuit may cause AKI through oxidative/nitrosative stress (26). Different from other variables, the shear stress perhaps directly related to circuit pressures, the impaired renal function caused by ECMO induced haemolysis would be magnified by prolongation of ECMO (26).

Evidence is growing that impaired function of one organ is communicated to the dysfunction of other organs and mediated via complex mechanisms in critical illness named “organ crosstalk” (38,39), AKI is one of a typical example involving in multiple organ interactions. Many systemic conditions such as vasculitis on ECMO patients invade both the liver and the kidney together, cause simultaneous liver and kidney injuries in both organs, additionally kidney may receive extra feedback from inflammation cytokine release due to decompensated cirrhosis and lead to subsequent renal vasoconstriction or structural kidney damage (38,39,40). AKI in liver cirrhosis patients is related to the abnormal systemic hemodynamics, splanchnic arterial vasodilatation and extra-hepatic vasoconstriction (40), ECMO implantation potentially precipitate as an acute event to further exaggerate already disturbed hemodynamics in advanced cirrhosis. Recent research reported that the most common precipitant of renal dysfunction in liver cirrhosis is bacterial infections which could product inflammatory cytokines, interfere the glomerular microcirculation or directly cause renal tubular cell death (41), which is a common incident on ECMO patients. Thus, these maybe explanations about patients with liver cirrhosis frequently present with or act as independently risk factor of AKI and its presence regardless acute in onset or gradual in, could lead to a steady decline of renal function or chronic renal failure (40).

The study has several limitations. Firstly, there are few original studies to record (< 10) the characteristics of AKI adult patients receiving ECMO. Although all the study had record ECMO support time, three studies were still excluded due to interquartile range (IQR) reported instead of mean ± standard deviation (SD). These data were not transfered to mean ± SD simply according to publicated formula (41,42) since potentially variables not follow normal distribution existed or inaccurate conversion; due to the complexity of diagnosis, various unstandardized definitions and diagnostic criteria existed throughout the records, it is only successful to extract two studies to analysis liver cirrhosis of severe AKI patients on ECMO and hindered the advancement of this research. However, this meta-analysis was based on extremely sensitive literature investigation and not merged the data together simply without carefully compared definition among studies, therefore the conclusion of this study still provided certain evidence. Extra analysis could not performed (indications, blood flow rate, ECMO modes et al) due to limited data collection. Furthermore studies are required to find out more risk factors of AKI among adult patients underwent the ECMO treatment.

## 5. Conclusions

This research is first time to provide the evidence that the risk of severe AKI requiring RRT was higher in liver cirrhosis patients and patients developed AKI/severe AKI with a longer ECMO support duration by systemic analysis, indicated ECMO support duration and liver cirrhosis may act as risk factors of AKI in adult patients received ECMO. It is still need more studies to support the conclusion and to detect more AKI risk factors among ECMO patients.

## Data Availability

All data generated or analyzed during this study are included in this article.

